# Prevalence of short peer reviews in leading general medical journals: a study of peer-review length at The BMJ, PLOS Medicine, and BMC Medicine

**DOI:** 10.1101/2023.05.07.23289620

**Authors:** Pascal Geldsetzer, Markus Heemann, Pauli Tikka, Grace Wang, Marika Mae Cusick, Ali Lenjani, Nandita Krishnan

## Abstract

**Background:** High-quality peer reviews are often thought to be essential to ensuring the integrity of the scientific publication process but measuring peer review quality is challenging. Although imperfect, review word count could serve as a simple, objective metric of review quality. We aimed to examine the prevalence of very short reviews and how often they inform editorial decisions on research articles in leading general medical journals.

**Methods:** We compiled a data set of peer reviews from published full-length original research articles in The BMJ, BMC Medicine, and PLOS Medicine for the years 2003 to 2022. In our primary analyses, we used a threshold of <200 words to calculate the prevalence of very short reviews. In secondary analyses, we also used thresholds of <100 and <300 words. In addition to disaggregating results by journal and year, we plotted the proportion of articles for which the first editorial decision was made based on a set of peer reviews in which very short reviews constituted 100%, ≥50%, ≥33%, or ≥20% of the reviews.

**Results:** In this sample of 11,466 reviews corresponding to 4,038 published articles, the median review word count was 425 (Interquartile Range=253–675), and the mean was 520 (Standard Deviation=401). The overall prevalence of very short (<200 words) reviews was 17.1% [95% CI: 16.4%–17.8%]. Across the three journals, 20.9% [95% CI: 19.6%–22.2%] of initial editorial decisions were based on review sets containing ≥50% very short reviews. The prevalence of very short reviews and share of editorial decisions based on review sets containing ≥50% very short reviews was highest for BMC Medicine at 26.8% [95% CI: 25.1%–28.5%] and lowest for The BMJ at 7.3% [95% CI: 5.7%–8.9%].

**Conclusions:** A substantial proportion of initial editorial decisions for published articles in these three leading general medical journals was based on peer reviews of such short length that they were unlikely to be of high quality. Future research should determine whether monitoring peer review length is a useful approach to improving the quality of the peer review process and which interventions, such as incentives and norm-based interventions, are most effective in soliciting more detailed reviews.

## Introduction

For many decades, peer review has been the key process on which the scientific community has relied to ensure that the reporting of research findings meets minimum quality standards.^1^ Peer review has become an established part of the scientific publication process and is used by most journals worldwide.^2,3^ Despite its widespread use, several deficiencies of the peer review process have been noted, such as the high cost and time burden on researchers. Aczel et al. estimated that researchers spent a total of over 100 million hours on peer review in 2020 alone, which translates to a monetary value between USD 400 million and 1.5 billion.^4^

Another deficiency is the duration of the peer review process, which can lead to delays in disseminating research findings. A systematic review found that the time from submission to publication for biomedical journals ranged from a few months to almost two years.^5^ Publication delays are especially problematic when research findings have time-sensitive implications for clinical guidelines and public health policies, as recently highlighted by the COVID-19 pandemic. Notwithstanding these deficiencies, a majority of researchers consider peer review important to ensure the quality and integrity of science.^6^

The effectiveness of peer review is contingent on its quality; however, it is unclear how prevalent low-quality reviews are and to what extent they inform editorial decisions on research articles. Likely reasons for the paucity of research in this area are the difficulty of measuring peer review quality and the lack of consensus on what constitutes quality.^6^ To date, the most commonly used approaches to evaluate peer review quality are editor and review quality evaluations.^7^ These typically consist of one or more items rated using Likert scales. Several scales have been developed but their validity has not been established.^7^ Additionally, these evaluations place an added burden on editors.^6^

Review volume (i.e., word count) could be a simple, objective metric for assessing peer review quality. A common recommendation in guides for conducting peer review is to provide comments that are sufficiently detailed.^8,9^ This is based on the rationale that a minimum number of words is required to provide constructive feedback. Review volume is certainly not a perfect measure of peer review quality. It is, after all, possible that a manuscript is of such outstanding quality that it cannot be further improved, such that a reviewer may not feel the need to provide more detailed comments. Conversely, a review that is long may not necessarily be a high-quality review if it lacks substantive content. Nevertheless, our study is based on the premise that whether or not a review is very short holds some valuable information in reflecting review quality. This premise is supported by research by Yadav et al., who found that review volume was strongly positively correlated with overall review quality.^10^ Additionally, a recent preprint study found that lengthier reviews were positively associated with the number of citations received by the article (arguably a measure of article quality).^11^

Thus, under the assumption that a very low word count reflects at least to some degree the quality of a peer review, this study aimed to determine the prevalence of very short peer reviews in three leading general medical journals – The BMJ, PLOS Medicine, and BMC Medicine. We defined very short reviews as having <200 words in our primary analyses, but – because there is no objective criterion to define such a word count threshold – also used additional cutoffs of <100 and <300 words in secondary analyses. We further aimed to determine the proportion of published research articles in these three journals for which initial editorial decisions were made based on very short peer reviews. We focused on leading general medical journals, reasoning that if high-tier journals must often rely on low-quality reviews, lower-tier journals are likely to do so even more frequently.

## Methods

### Data sources

We selected all journals in the “Medicine, General & Internal” category of Clarivate’s Journal Citation Reports that were among the 20 journals with the highest 2020 Journal Impact Factor and have published peer review reports of at least a subset of full-length research articles that have appeared in the journal.^12^ We used the 2020 Journal Impact Factor to avoid the possibly transient effects of the COVID-19 pandemic on journal rankings by impact factor. Three journals met these criteria: The BMJ, BMC Medicine, and PLOS Medicine. While The BMJ has a fully open peer review process that publishes the peer review history and reviewer names by default, BMC Medicine runs a peer review process in which peer review history is published, but reviewers are given the choice to disclose their identity. PLOS Medicine lets authors opt-in to publish the peer review history of their accepted manuscript and only discloses reviewer names if the reviewer chooses to sign their review. All three journals publish the pre-publication history of accepted manuscripts, which includes all previous versions of the manuscript, reviewer comments at each stage, and authors’ responses to comments from reviewers and editors.^13,14^ Only The BMJ provides the self-reported job title of the reviewer.

The BMJ began publishing peer reviews in 2015, BMC Medicine in 2003, and PLOS Medicine in 2019. The most recent impact factors (2021) of these journals were: 96.21 (The BMJ), 11.81 (BMC Medicine), and 11.61 (PLOS Medicine).^15^ Our analysis covered peer reviews published through December 2022 for all journals. We restricted our analysis to peer reviews of full-length original research articles as word count cutoffs for defining quality may be different for peer reviews of shorter research works, such as brief reports. Because none of the three journals publish reviews for articles that were outright rejected, our sample consisted only of reviews for articles that were eventually published. Only the first set of peer reviews that an article received (i.e., reviews used to arrive at an initial editorial decision of accept or revise and resubmit) was included in the analysis. The three journals did not openly make available peer reviews for all research articles published; however, if peer reviews for an article were made available openly, all (rather than a subset) peer reviews that the article received were made available. We developed code in Python to count the words of each published peer review. The scraping of peer review reports from the journal websites was carried out between 25^th^ November 2022 and January 27^th^ 2023.

### Definition of peer review quality

In our primary analysis, we defined very short peer reviews as those containing <200 words. Although there is no objective criterion to inform the selection of a word count threshold that best reflects peer review quality, we chose the cutoff of <200 words for the following reasons. Given that the three journals have abstract lengths between 250-350 words, we believe that reviews that are significantly shorter than a high-level manuscript summary are unlikely to provide sufficiently detailed feedback. Additionally, established instruments for measuring the quality of peer review, such as the JBJS-Peer Review scoring scale, have used 200 words as the minimum word count threshold that corresponds to the highest scoring category.^16^ Nevertheless, we recognize that there is some subjectivity to selecting this cutoff and therefore, also present results using cutoffs of <100 and <300 words.

### Statistical analysis

We created plots to examine the distribution of the number of words across peer reviews and the number of reviews across research articles for all journals and years combined. We also calculated the distribution (mean and median) of the number of reviews per article and words per review disaggregated by journal and year. We used the Kruskal-Wallis test to assess significant differences in median review word counts across journals. To test for significant differences in mean review word counts across journals, we used one-way ANOVA with the Holm’s method to adjust for multiple comparisons.^17^ Following this, we categorized word counts of peer reviews (0-99; 100-199; 200-299; 300-399; 400- 499; 500+) and calculated the prevalence of reviews for each word count category by year for each journal.

Given the importance of the first editorial decision, we analyzed the proportion of articles for which this initial decision was based on very short peer reviews (i.e., reviews of <200 words). For each article, we categorized its associated peer reviews into one of four groups (that were not mutually exclusive) based on the proportion of very short reviews in the set. A set was classified as containing 100% very short reviews (i.e., all reviews in the set were <200 words); ≥50% very short reviews (i.e., at least half of the reviews in the set were <200 words); ≥33% very short reviews (i.e., at least a third of reviews in the set were <200 words); and ≥20% very short reviews (i.e., at least a fifth of reviews in the set were <200 words). We plotted the proportion of articles for which the first editorial decision was made based on the proportion of very short reviews in the set for each journal and for all journals combined. We repeated the same analyses using word count cutoffs of <100 and <300 words.

Finally, we also plotted the distribution of peer review word count by reviewer seniority for reviews from The BMJ. We defined senior scientists as those whose self-reported job title contained any of the following terms: professor (excluding those with the addition of “assistant” or associate”), dean, head, chief, director or chair. All other reviewers were classified as junior scientists.

## Results

Our dataset consisted of 11,466 reviews corresponding to 4,038 articles. The distribution of the number of peer reviews per article showed that a little less than half (42.7% [95% CI: 41.2% – 44.2%]) of all included articles received two reviews (Figure 1). A notable exception was PLOS Medicine, where almost half (49.9% [95% CI: 45.0% – 54.7%]) of all articles received four initial reviews (Figure S1). Regarding the distribution of the number of words per review, the highest proportion of reviews had approximately 250 words (Figure 1). The distribution was right skewed, with 5.9% of reviews containing <100 words; 17.1% containing <200 words; 72.7% containing between 200 and 1,000 words; and few (10.2%) containing >1,000 words. Distributions of word counts per peer review and reviews per research article by journal and year are shown in the Supplementary Material (Figures S1, S2, S3, S4).

**Figure 1.**
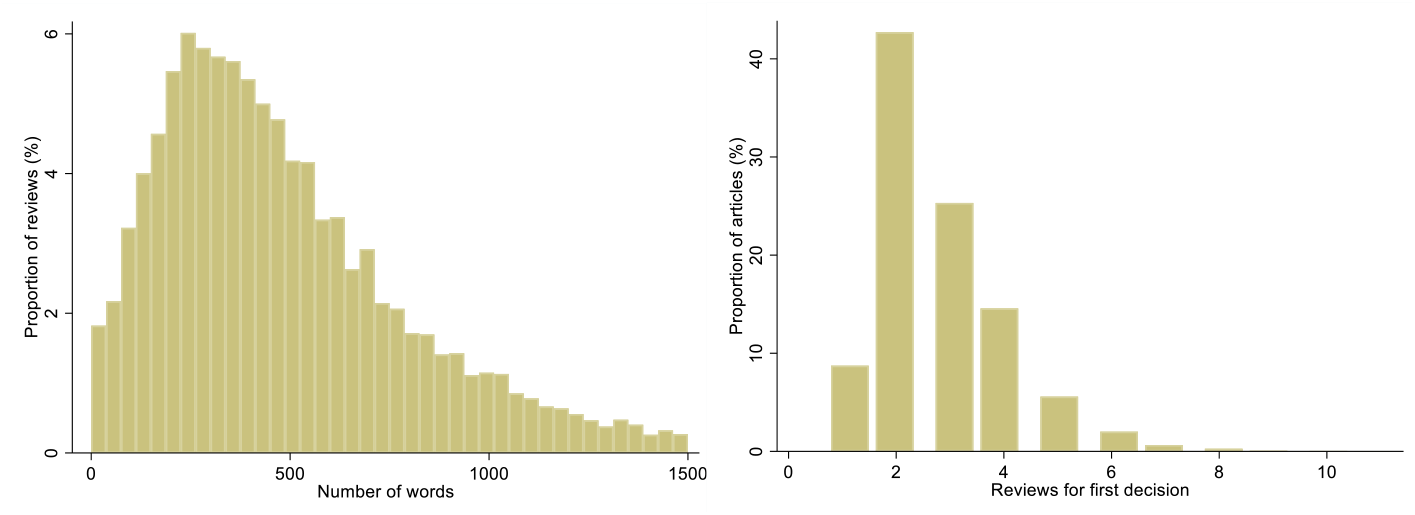
Distribution of the number of words per peer review and the number of peer reviews per research article for all years and journals combined. **Notes**. Reviews with a word count >1,500 (325 of 11,466 reviews [2.8%]) are not shown. No articles received more than nine peer reviews.

The distribution of the number of reviews per article and word count per review disaggregated by journal and year are shown in Table 1. Across all three journals and all years analyzed, the median word count per review was 425 (Interquartile Range (IQR): 253–675), and the mean was 520 (SD=401). The BMJ had the highest median and mean number of words per review, at 525 (IQR: 318–820) and 636.0 (SD=464.4) words, respectively. BMC Medicine had the lowest median and mean number of words per review, at 376 (IQR: 221–590) and 455.2 (SD=350.2) words, respectively. The Kruskal-Wallis test showed significant differences in the median review word count across all three journals (p<0.001). The F-test for any difference in mean peer review word count across the three journals was also significant (p<0.001). In pairwise testing, differences in the mean word count between all pairs of journals were statistically significant (p<0.001).

**Table 1.** Distribution of the number of peer reviews per research article and word count per peer review by journal and year. **Notes**. “NA” indicates that no open peer reviews were available for the given journal in that year. Only the first set of peer reviews that an article received (i.e., the peer reviews used to arrive at a first editorial decision of accept or revise and resubmit) was included in the analysis. The analysis was restricted to research articles only. ^1^The three journals did not openly make available peer reviews for all research articles published; however, if peer reviews for an article were made available openly, all (rather than a subset) peer reviews that the article received were made available.

| *Year* | *# of reviews* | *Median # of reviews per article (IQR)<sup>†</sup>* | *Median # of words per review (IQR)* | *# of reviews* | *Median # of reviews per article (IQR)<sup>†</sup>* | *Median # of words per review (IQR)* | *# of reviews* | *Median # of reviews per article (IQR)<sup>†</sup>* | *Median # of words per review (IQR)* | *# of reviews* | *Median # of reviews per article (IQR)<sup>†</sup>* | *Median # of words per review (IQR)* |
| --- | --- | --- | --- | --- | --- | --- | --- | --- | --- | --- | --- | --- |
|  | **All Journals** |  |  | **BMC Medicine** |  |  | **The BMJ** |  |  | **PLOS Medicine** |  |  |
| All years | 11466 | 3 (2, 3) | 425 (253, 675) | 6086 | 2 (2, 3) | 376 (221, 590) | 3816 | 4 (3, 5) | 525 (318, 820) | 1564 | 4 (3, 4) | 411 (254, 652.5) |
| 2003 | 4 | 2 (2, 2) | 416 (206.5, 1048) | 4 | 2 (2, 2) | 416 (206.5, 1048) | NA | NA | NA | NA | NA | NA |
| 2004 | 65 | 2 (2, 2) | 393 (199, 580) | 65 | 2 (2, 2) | 393 (199, 580) | NA | NA | NA | NA | NA | NA |
| 2005 | 28 | 2 (1, 3) | 270 (156.5, 421) | 28 | 2 (1, 3) | 270 (156.5, 421) | NA | NA | NA | NA | NA | NA |
| 2006 | 60 | 2 (2, 3) | 357 (256, 575) | 60 | 2 (2, 3) | 357 (256, 575) | NA | NA | NA | NA | NA | NA |
| 2007 | 71 | 2 (2, 3) | 438 (256, 645) | 71 | 2 (2, 3) | 438 (256, 645) | NA | NA | NA | NA | NA | NA |
| 2008 | 37 | 2 (2, 3) | 325 (240, 580) | 37 | 2 (2, 3) | 325 (240, 580) | NA | NA | NA | NA | NA | NA |
| 2009 | 33 | 2 (2, 2) | 380 (232, 520) | 33 | 2 (2, 2) | 380 (232, 520) | NA | NA | NA | NA | NA | NA |
| 2010 | 70 | 2 (2, 3) | 290 (165, 559) | 70 | 2 (2, 3) | 290 (165, 559) | NA | NA | NA | NA | NA | NA |
| 2011 | 148 | 2 (2, 3) | 316 (180.5, 501.5) | 148 | 2 (2, 3) | 316 (180.5, 501.5) | NA | NA | NA | NA | NA | NA |
| 2012 | 221 | 2 (2, 3) | 282 (145, 470) | 221 | 2 (2, 3) | 282 (145, 470) | NA | NA | NA | NA | NA | NA |
| 2013 | 333 | 2 (2, 3) | 313 (172, 529) | 333 | 2 (2, 3) | 313 (172, 529) | NA | NA | NA | NA | NA | NA |
| 2014 | 313 | 2 (2, 2) | 335 (198, 563) | 313 | 2 (2, 2) | 335 (198, 563) | NA | NA | NA | NA | NA | NA |
| 2015 | 671 | 2 (2, 3) | 406 (240, 683) | 380 | 2 (2, 2) | 345.5 (195.5, 582.5) | 291 | 3 (2, 4) | 482 (309, 762) | NA | NA | NA |
| 2016 | 679 | 2 (1, 3) | 538 (336, 819) | 204 | 1 (1, 2) | 404 (211.5, 628.5) | 475 | 3 (2, 4) | 576 (389, 978) | NA | NA | NA |
| 2017 | 876 | 3 (2, 3) | 452.5 (253.5, 725.5) | 443 | 2 (2, 3) | 368 (212, 591) | 433 | 3 (3, 4) | 566 (314, 862) | NA | NA | NA |
| 2018 | 979 | 3 (2, 3) | 436 (262, 694) | 505 | 2 (2, 3) | 391 (230, 617) | 474 | 3 (3, 4) | 488 (317, 783) | NA | NA | NA |
| 2019 | 1052 | 3 (2, 3) | 444.5 (270.5, 677.5) | 486 | 2 (2, 3) | 411.5 (260, 605) | 463 | 3 (2, 4) | 509 (298, 773) | 103 | 4 (4, 4) | 381 (241, 621) |
| 2020 | 1824 | 3 (2, 4) | 410 (239, 683) | 828 | 2 (2, 3) | 370.5 (217.5, 618) | 642 | 4 (3, 5) | 531.5 (299, 794) | 354 | 4 (1, 4) | 358 (191, 557) |
| 2021 | 1941 | 3 (2, 4) | 432 (273, 670) | 751 | 3 (2, 3) | 402 (249, 579) | 527 | 4 (3, 5) | 508 (308, 798) | 663 | 4 (4, 4) | 417 (274, 672) |
| 2022 | 2061 | 3 (2, 4) | 440 (277, 678) | 1106 | 2 (2, 3) | 411 (261, 611) | 511 | 4 (3, 5) | 517 (317, 865) | 444 | 4 (4, 5) | 452.5 (281, 689) |

Figure 2 shows the proportion of articles for different review word count categories by year for each journal. For all journals, peer review word counts remained relatively consistent across time. Of the three journals, BMC Medicine had the greatest proportion of overall reviews with <200 words (21.0% [95% CI: 19.9%–22.0%]) (see also Figure S1). The BMJ had the lowest proportion of reviews with <200 words (10.9% [95% CI: 9.9%–11.9%]).

**Figure 2.**
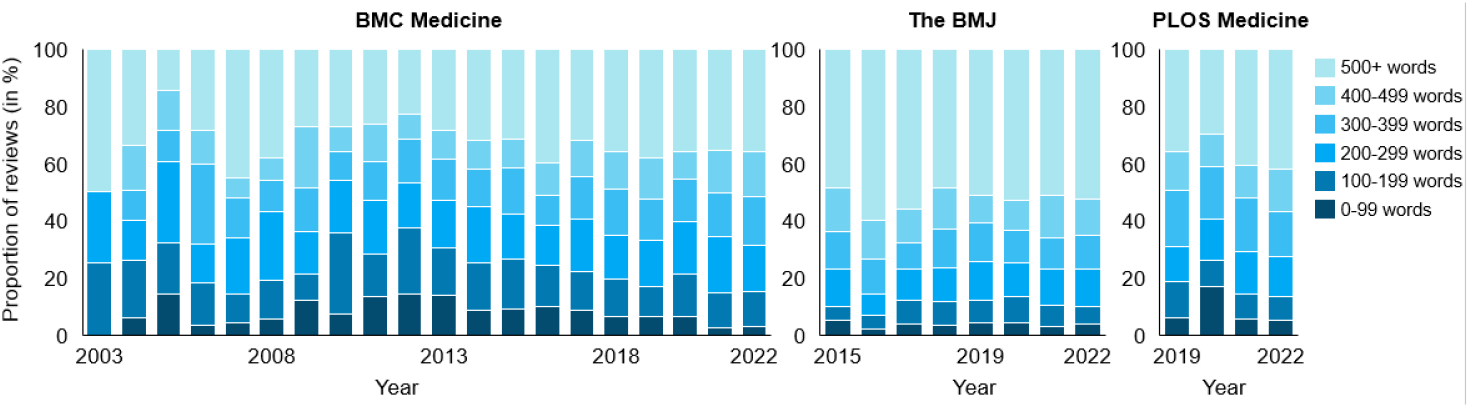
The proportion of peer reviews in different word count categories by journal and year.

Comparing the share of very short reviews per article by applying equal article weights rather than equal review weights yielded similar results. Across all journals, 18.6% (95% CI: 17.7%–19.5%) of reviews per article contained <200 words. This proportion was highest among PLOS Medicine articles, where 23.9% (95% CI: 20.8%–27.0%) of reviews per article were <200 words. For BMC Medicine and The BMJ, 20.9% (95% CI: 19.8%–22.1%) and 10.7% (95% CI: 9.6%–11.8%) of reviews contained <200 words, respectively.

Figure 3 shows the proportion of articles for which the first editorial decision was made based on a set of peer reviews that consisted of 100%, ≥50%, ≥33%, or ≥20% very short reviews. Across journals, 20.9% of first editorial decisions were based on review sets containing ≥50% very short reviews. BMC Medicine had the highest proportion of initial editorial decisions that were based on peer review sets in which half or more of the reviews were very short (26.8% [95% CI: 25.1%–28.5%]). The BMJ had the lowest proportion of editorial decisions based on peer review sets with ≥50% very short reviews (7.3% [95% CI: 5.7%-8.9%]). Similar trends were observed when we used cutoffs of <100 words (Figure S5) and <300 words (Figure S6) to define very short reviews.

**Figure 3.**
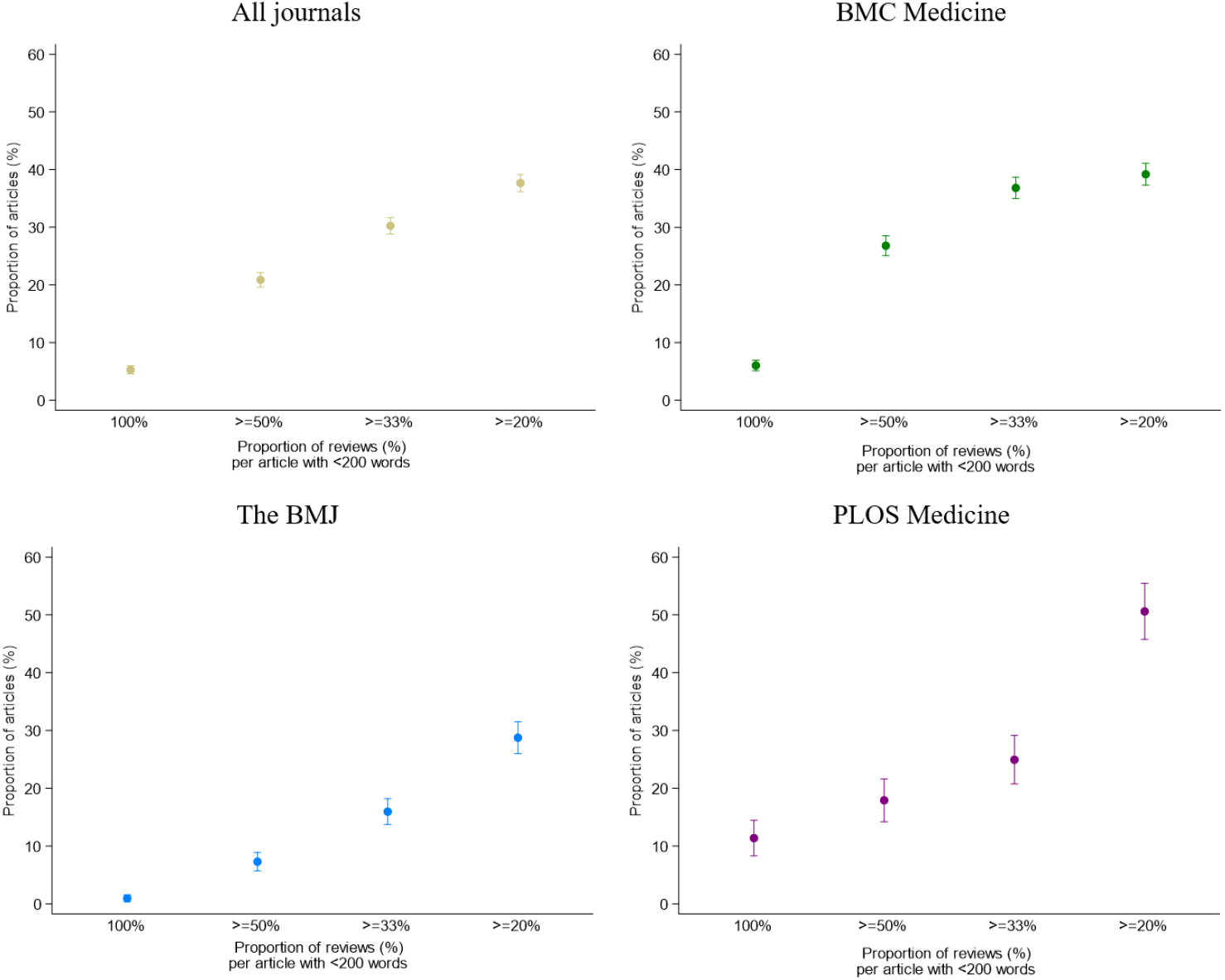
Proportion of articles for which the first editorial decision was based on peer review sets consisting of 100%, ≥50%, ≥33%, or ≥20% very short (<200 words) reviews. **Notes**. The denominator for the proportion of articles is the total number of research articles that were published and for which the journal made all peer reviews openly available.

Lastly, we analyzed the distribution of the number of words per peer review by reviewer seniority for The BMJ. The mean number of words per review was similar for senior (631.5 words [95% CI: 611.8– 651.2]) and junior (643.2 words [95% CI: 618.3–668.1]) scientists. The proportion of reviews in different word count categories was also similar for senior and junior scientists. Specifically, for senior scientists, 10.1% (95% CI: 8.4%–11.8%) of reviews had <200 words; 72.3% (95% CI: 69.8%–74.8%) had 200–1000 words; and 17.6% (95% CI: 15.4%–19.7%) had >1000 words (Figure 4). For junior scientists, the corresponding percentages were 11.4% (95% CI: 10.1%–12.7%) for reviews with <200 words, 72.2% (95% CI: 70.3%–74.0%) for reviews between 200–1000 words, and 16.4% (95% CI: 14.9%–17.9%) for reviews >1000 words (Figure 4).

**Figure 4.**
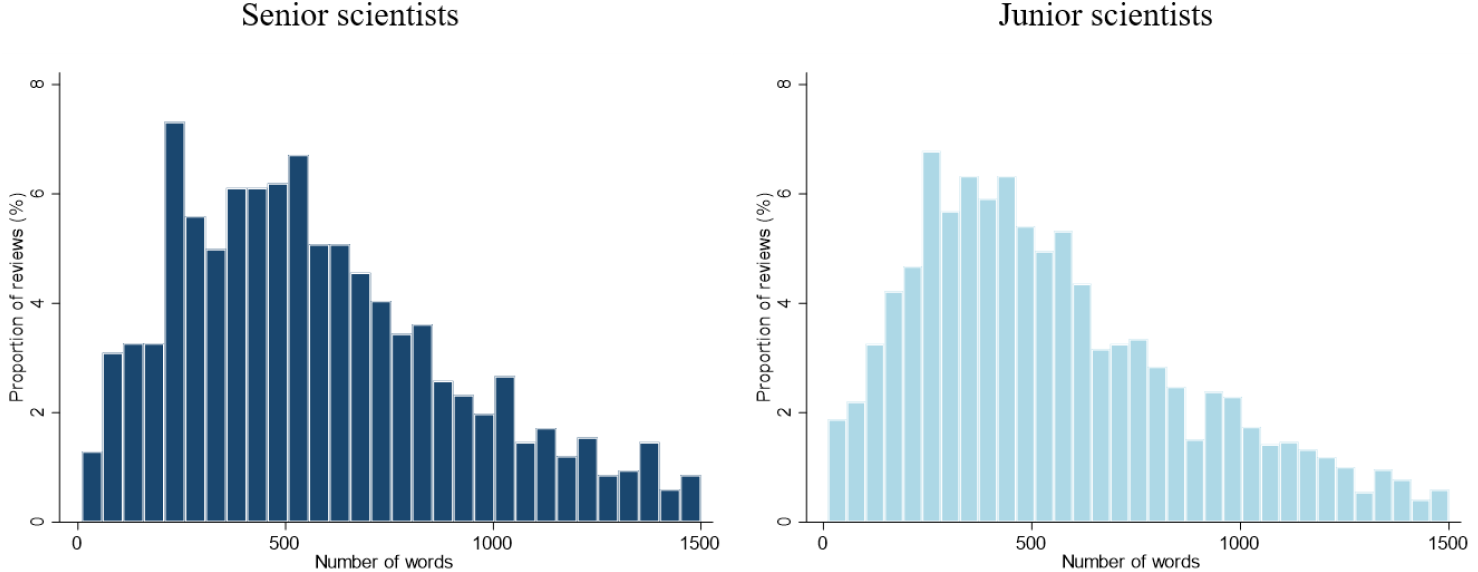
Distribution of the number of words per peer review in The BMJ by reviewer seniority. **Notes**. This analysis was restricted to The BMJ (n=3,817 peer reviews) because this was the only journal that made the self-reported job title of the peer reviewer available. For 7.7% of reviews (n=292), the job titles of the reviewers were missing. Senior scientists (n=1,218) were defined as those self-reporting a job title containing any of the following terms: professor (without the addition of the words “assistant” or “associate”), dean, head, chief, director, or chair. Reviewers with all other job titles were considered junior scientists (n=2,305). Reviews with more than 1,500 words (n=181) were excluded in this figure.

## Discussion

In this analysis of peer reviews from three leading medical journals, we found that – across all journals and years – 17% of reviews were very short (i.e., <200 words). We also found that one in five initial editorial decisions that eventually led to publication were based on review sets in which very short reviews accounted for half or more of the reviews. Even when using a more conservative threshold (<100 words) to define very short reviews, around one in 20 articles were published based on an initial editorial decision that had to rely on a peer review set in which such excessively short reviews constituted at least half of the reviews.

We identified some variation in these findings across journals. Relative to PLOS Medicine and BMC Medicine, The BMJ had significantly longer reviews on average and a lower share of very short reviews. Additionally, The BMJ had the lowest proportion of editorial decisions made based on peer review sets with very short reviews. These results were consistent across word count thresholds used to define very short reviews. Review word count has been shown to be positively correlated with journal impact factor;^6,18-19^ thus, these findings may be related to differences in journal impact factors (The BMJ: 96.21; vs. PLOS Medicine: 11.61 and BMC Medicine: 11.81). However, the mechanisms underlying this association are not fully clear. It is possible that reviewers invest more effort when conducting reviews for higher impact journals.^6,19^ Similarly, it is possible that higher impact journals are able to identify willing reviewers that, on average, provide more thorough reviews.^6^ We did not find significant differences in review word count by review seniority among reviews for The BMJ. This is consistent with previous research, which found that reviewer seniority was not a significant predictor of review quality.^20^ However, other reviewer characteristics for which we did not have data may also be relevant. For instance, some data suggest that reviewers from predominantly English-speaking countries tend to write longer reviews, and it is possible that higher impact journals are more likely to solicit reviewers from these countries.^6,19^

The differences in the prevalence of very short peer reviews across the three journals in our study could also stem from differences in the design of the peer review process itself. Of the three journals examined, only The BMJ has a completely open peer review process, in which the reviews and names of reviewers are published by default without the ability to opt-out. Additionally, based on information provided by each of the journals on their websites, it appears that The BMJ provides the most incentives (all non- monetary) for reviewers (e.g., APC discounts, CME credits, reviewer certificates).^21-23^ Such incentives could constitute a motivation to perform the reviews more diligently. It is, thus, possible that increasing accountability and providing incentives are effective approaches to encourage reviewers to provide more detailed reviews.^24,25^ Norm-based interventions have a strong evidence base for changing behaviors across various contexts and could also potentially be effective in this context.^26^ Future studies could investigate the impact on review word count of interventions that target norms, such as posting sample reviews and indicating to reviewers how the word count of their review compares on average to other reviews for similar articles.^27^

We reiterate that using word count as a measure of review quality has limitations. A review could be very short because the article had no flaws that merited critiquing. Similarly, a longer review may not necessarily be of higher quality if it does not provide competent feedback. Thus, other attributes of a review, such as its relevance and coverage, should ideally also be considered when assessing review quality.^27^ Nevertheless, word count appears to be an important component of quality^10^ and is a simple and easily computable metric. We also acknowledge that our selection of a cutoff of <200 words to define very short reviews is arbitrary. Recognizing this limitation, we show all results using alternative cutoffs.

An additional limitation of our study is that we only had access to the review history for those articles that were eventually published. Furthermore, in the case of PLOS medicine, authors had to actively opt-in to make the review history for their article publicly available. Thus, it is unclear whether our findings generalize to reviews of articles that were rejected or for which reviews were not made available. We also only analyzed data from three medical journals, which is a small sample. However, given that these three journals are high-impact general medical journals, we believe that our estimates are conservative for the medical field more generally based on the assumption that it is likely that lower-tier journals have a higher prevalence of very short reports and rely more frequently on these types of reports to make editorial decisions.

Our findings show a substantial prevalence of very short reviews in three leading general medical journals, such that roughly one in five editorial decisions are made based on review sets in which reviews of <200 words constitute half or more of the reviews. Future studies could investigate whether monitoring peer review length is a useful approach to improving the quality of the peer review process. Future research could also assess the effectiveness of different interventions, such as incentives and norm-based interventions, in eliciting more detailed reviews.

## Data Availability

All data produced in the present study are available upon reasonable request to the authors

## Author Contributions

Concept and design: Geldsetzer

Acquisition of Data: Lenjani, Tikka

Statistical analysis and interpretation of data: Geldsetzer, Heemann, Wang, Cusick, Krishnan

Drafting of the manuscript: Geldsetzer, Heemann, Wang, Krishnan

Critical revision of the manuscript for important intellectual content: Geldsetzer, Heemann, Tikka, Wang, Cusick, Lenjani, Krishnan

Supervision: Geldsetzer

