## Supplement Figures for "Prevalence of short peer reviews in leading general medical journals: a study of peer-review length at The BMJ, PLOS Medicine, and BMC Medicine"

### Supplementary Materials

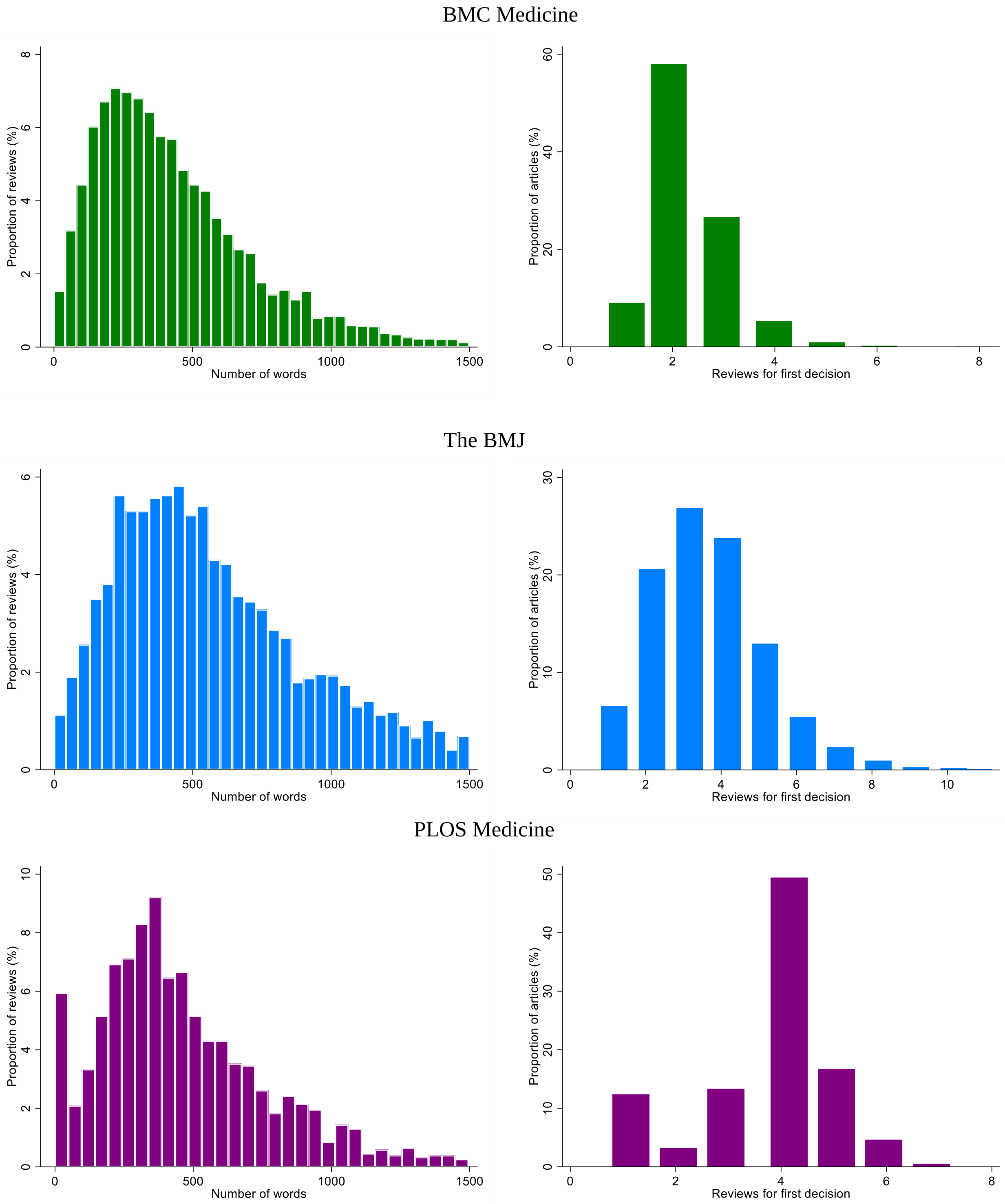


**Figure S1: Distribution of the number of words per peer review and the number of peer reviews per research article by journal**

The distribution of word counts per peer review (left column of panels) and reviews per research article (right column of panels) are shown for BMC Medicine (green), The BMJ (blue), and PLOS Medicine (purple) from 2003-2020. Reviews with over 1500 words were excluded.

# 2003-2004

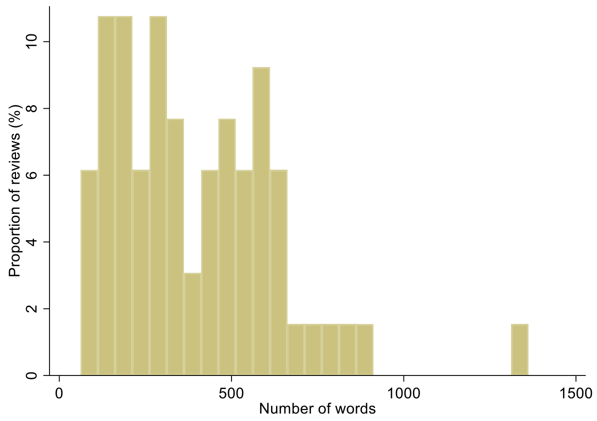


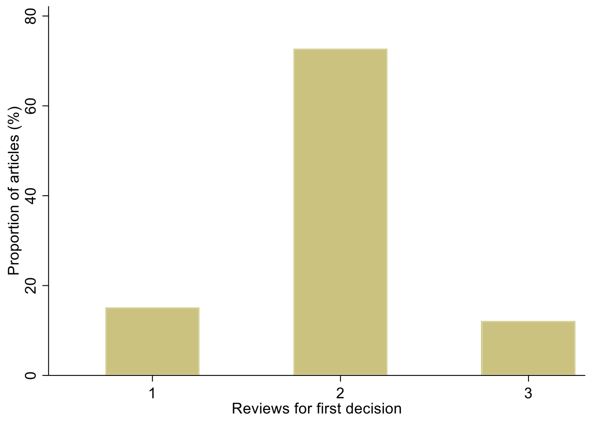


# 2005-2006

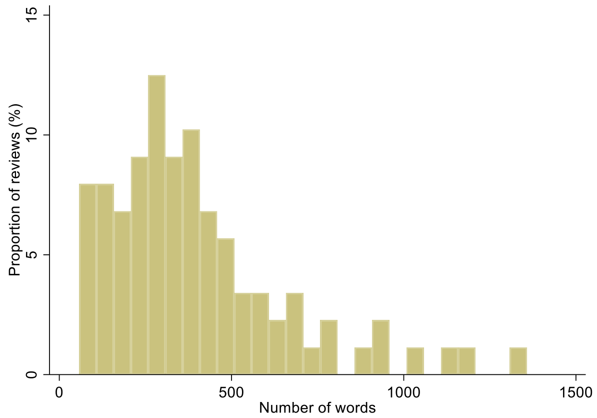


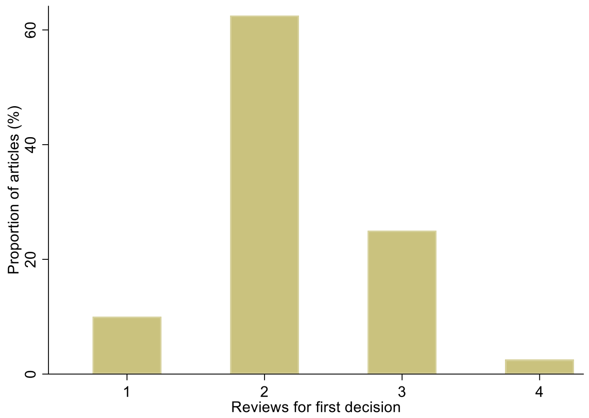


# 2007-2008

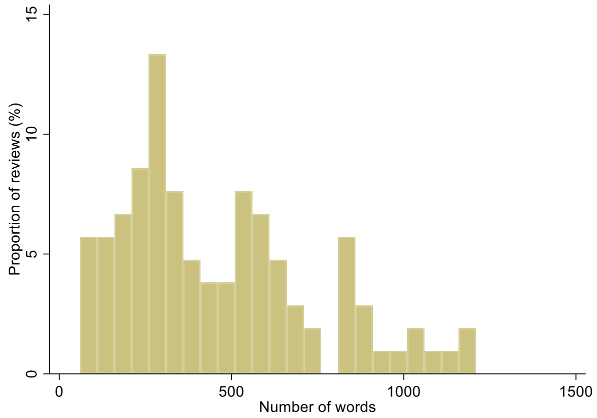


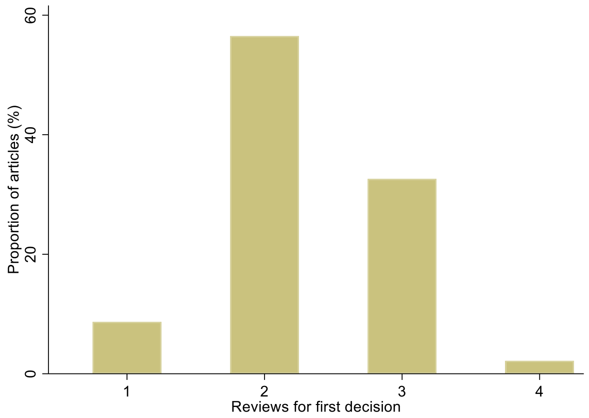


# 2009-2010

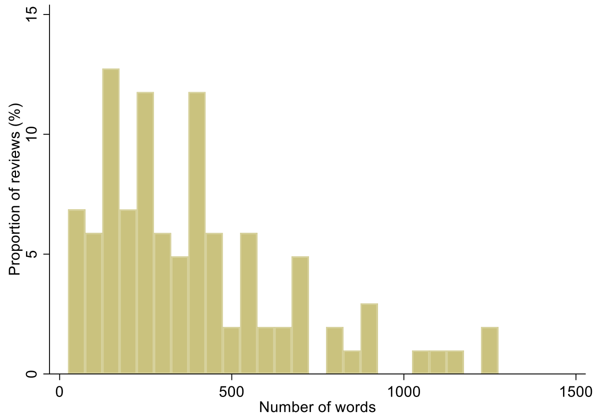


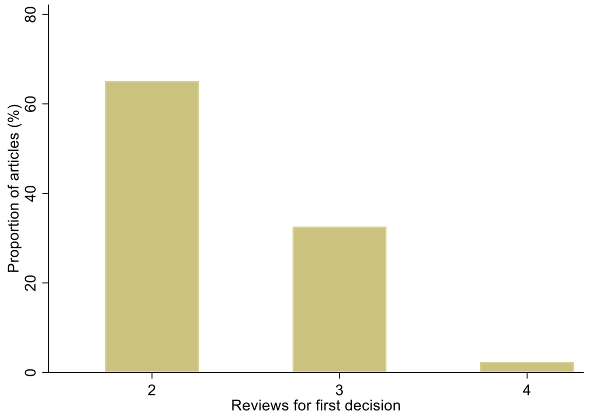

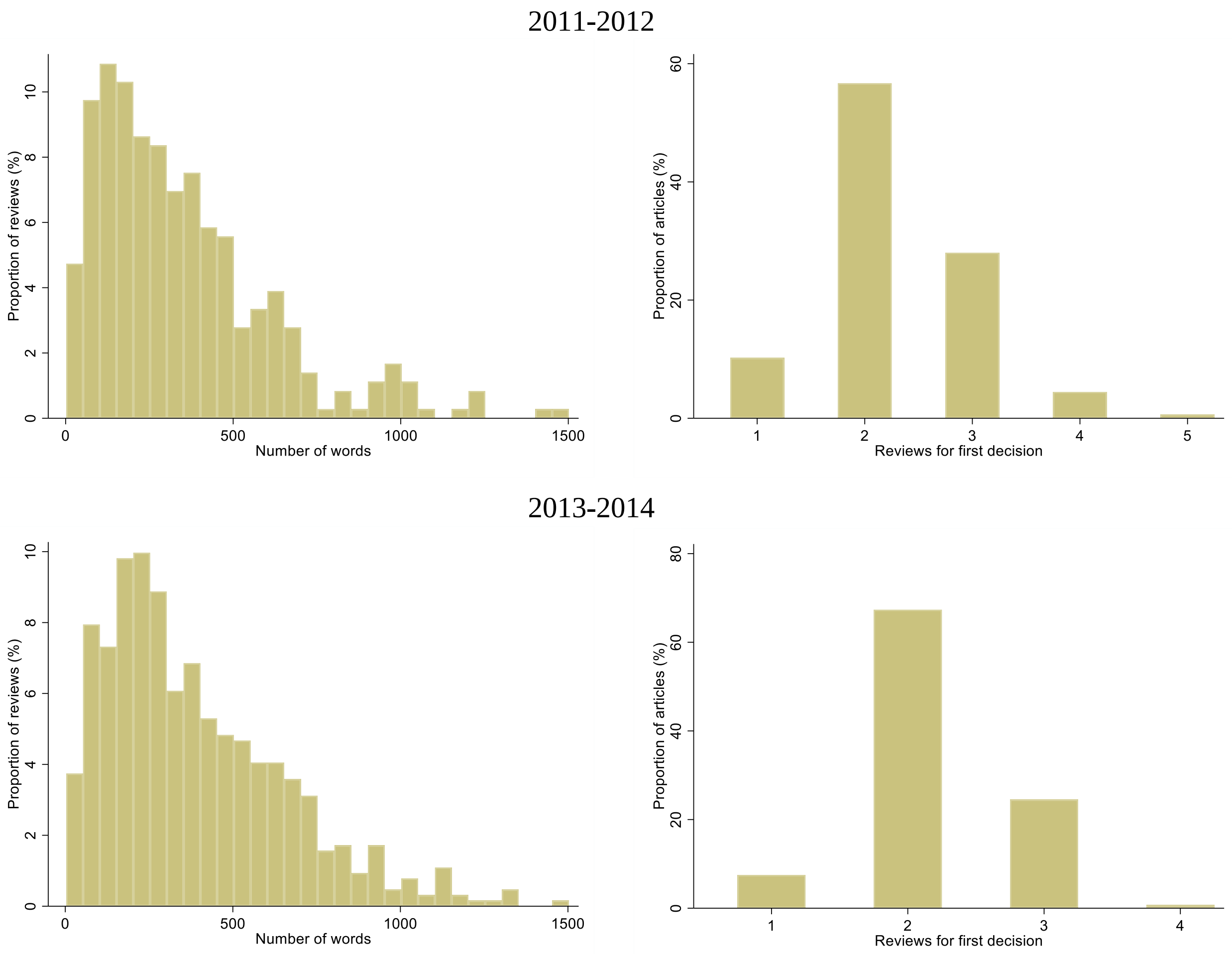


**Figure S2: Distribution of the number of words per peer review and the number of peer reviews per research article for BMC Medicine between 2003-2014.**

Reviews with over 1500 words were excluded.

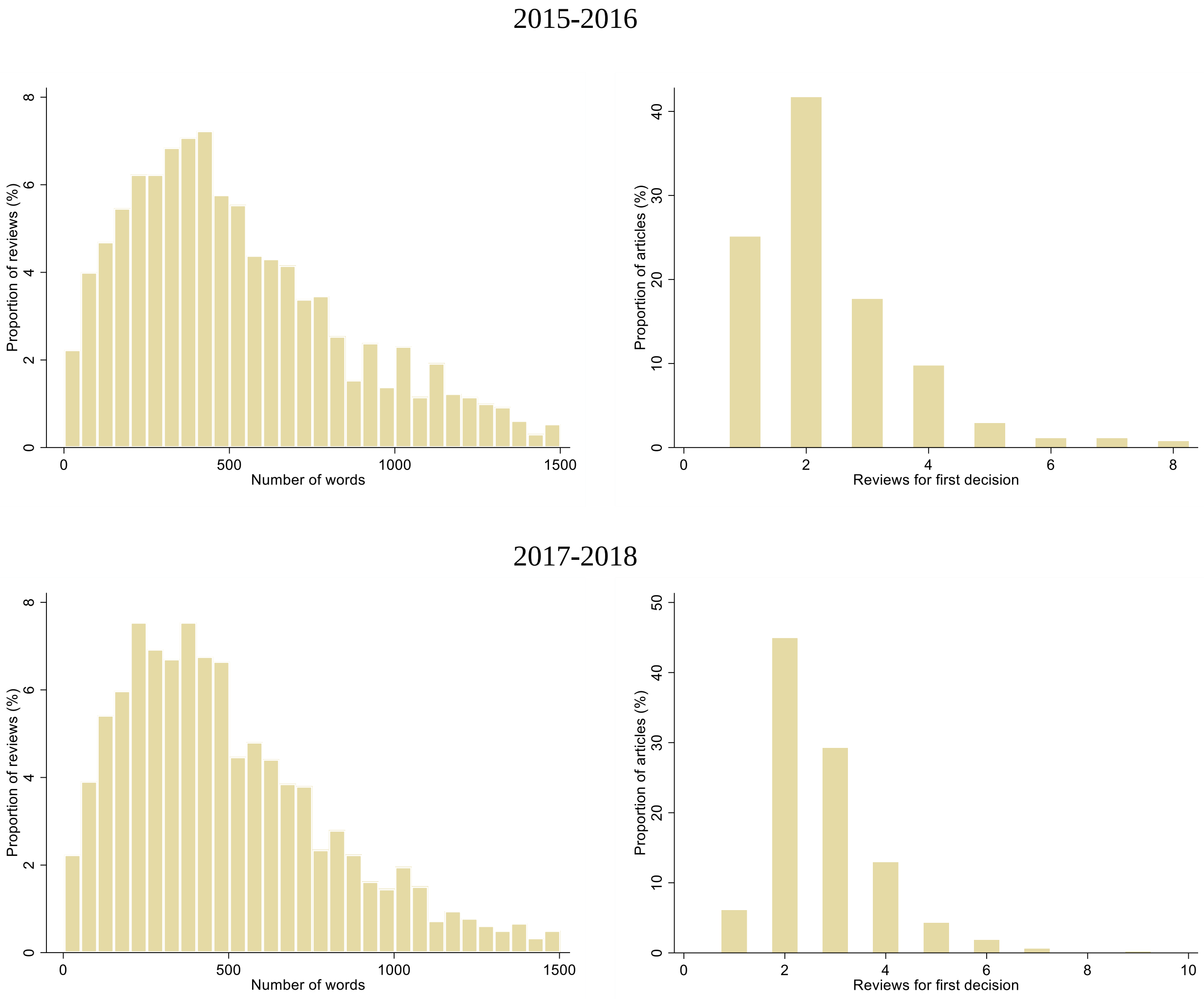


**Figure S3: Distribution of the number of words per peer review and the number of peer reviews per research article for BMC Medicine and The BMJ between 2015-2018.**

Reviews with over 1500 words were excluded.

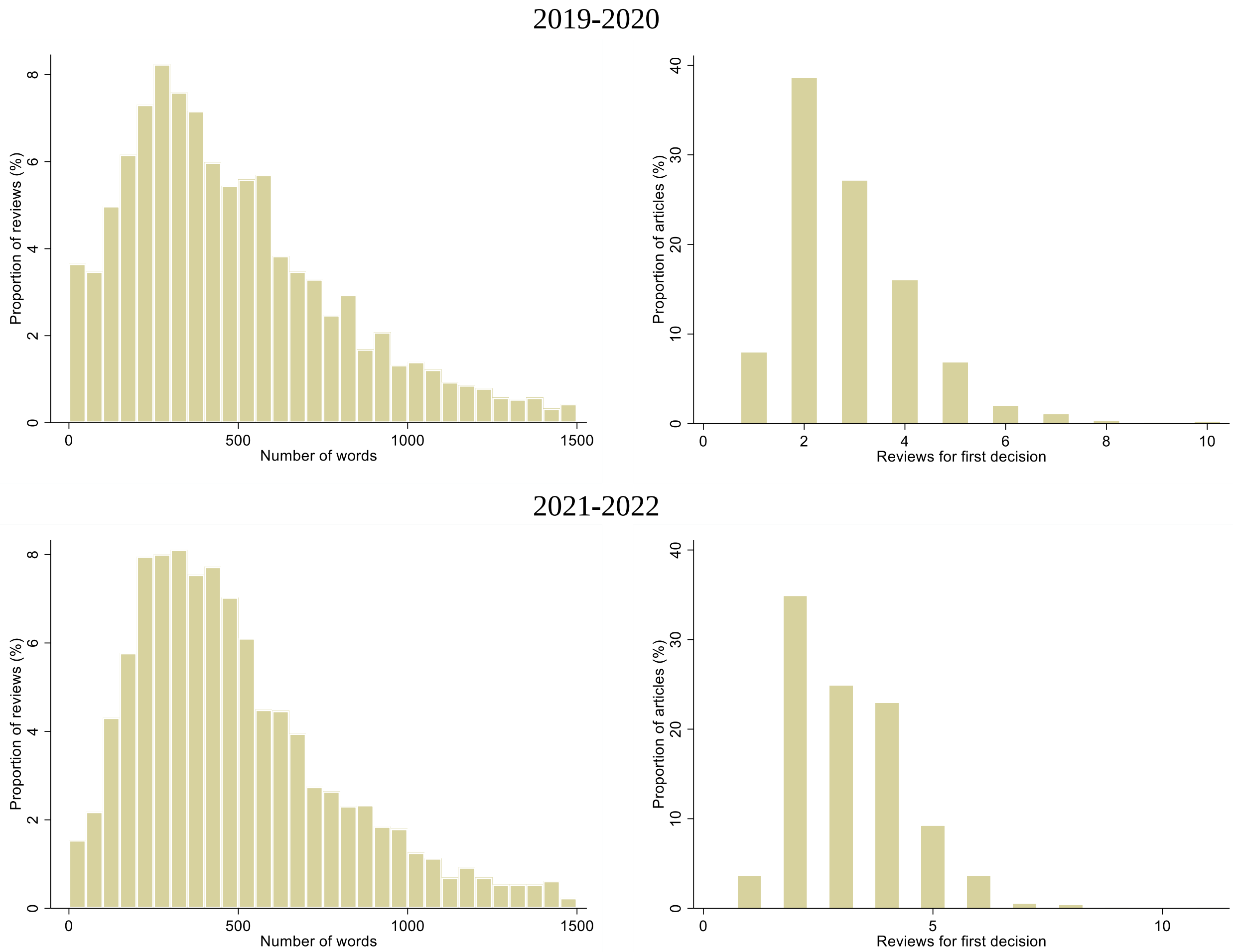


**Figure S4: Distribution of the number of words per peer review and the number of peer reviews per research article for BMC Medicine, The BMJ, and PLOS Medicine between 2019-2022.**

Reviews with over 1500 words were excluded.

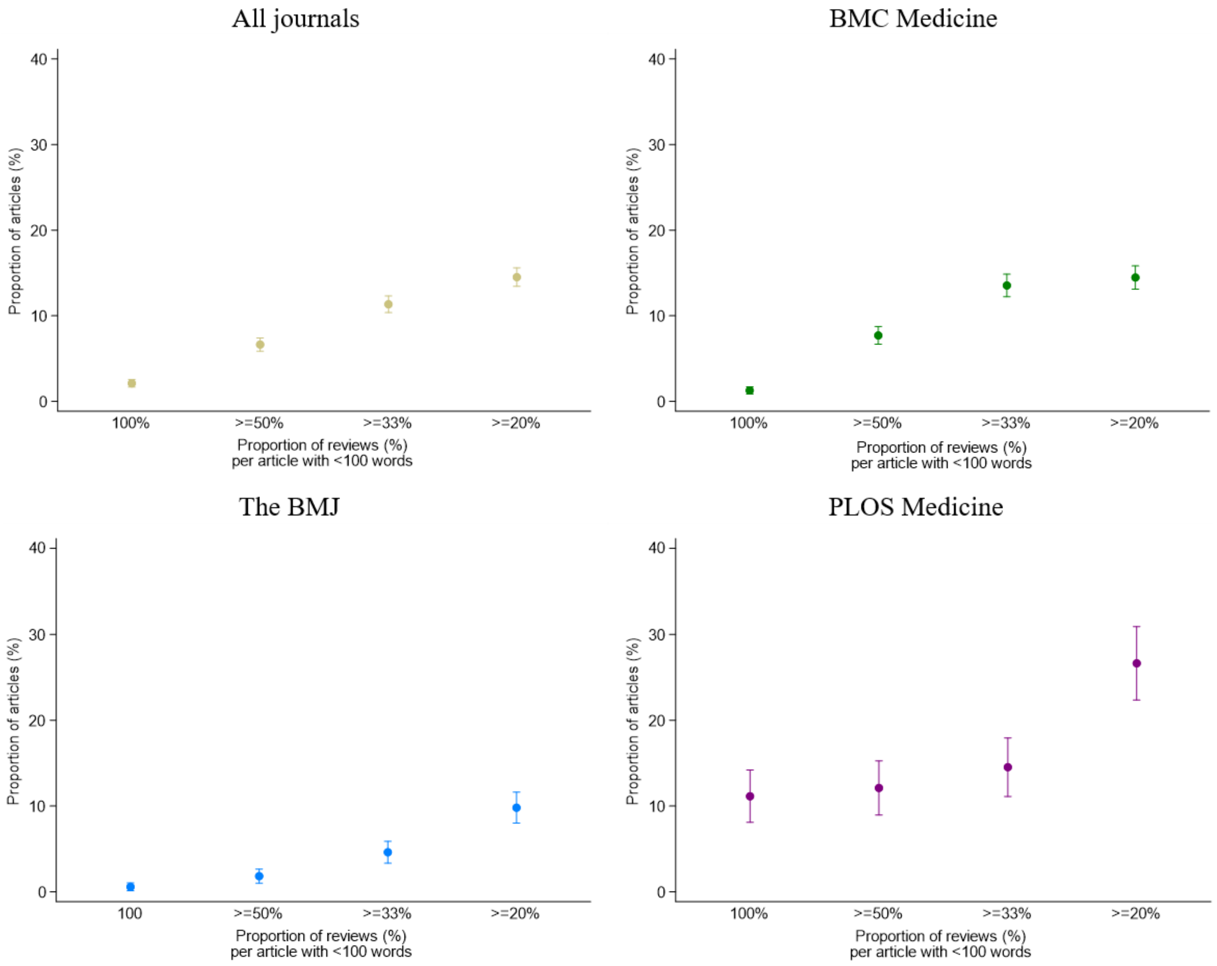


**Figure S5: Proportion of articles for which the first editorial decision was made based on a set of peer reviews consisting of 100%, ≥50%, ≥33%, or ≥20% of reviews with <100 words.**

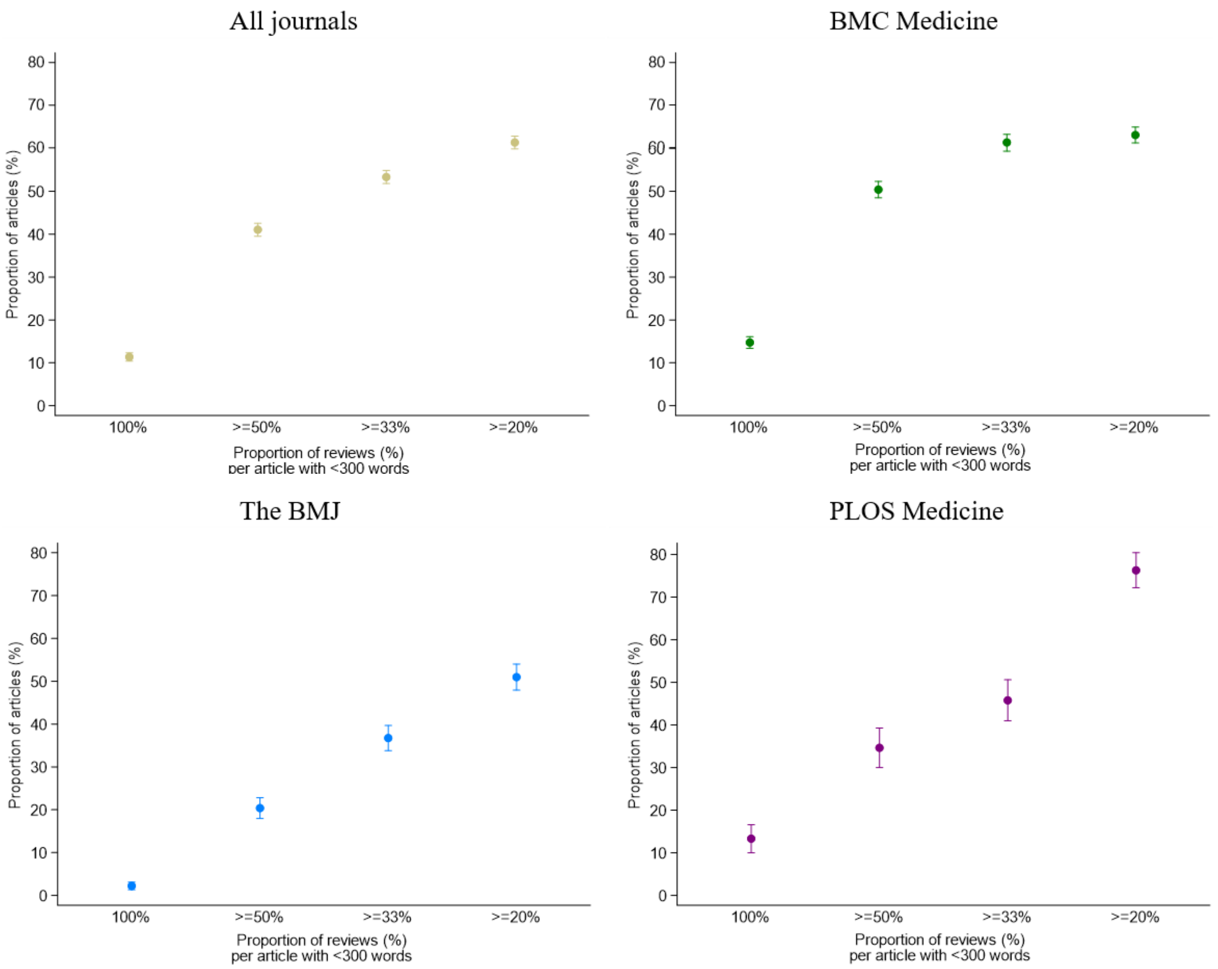


**Figure S6: Proportion of articles for which the first editorial decision was made based on a set of peer reviews consisting of 100%, ≥50%, ≥33%, or ≥20% of reviews with <300 words.**
